# Predictors for the Recurrence and Prognosis of Stroke patients within a Multi-Ethnic Asian Population

**DOI:** 10.1101/2023.09.01.23294970

**Authors:** Keng Siang Lee, Isabel Siow, Tessa Riandini, Kaavya Narasimhalu, Kelvin Bryan Tan, Deidre Anne De Silva

**Affiliations:** Department of Neurosurgery, King’s College Hospital, London, UK; Department of Basic and Clinical Neurosciences, Maurice Wohl Clinical Neuroscience Institute, Institute of Psychiatry, Psychology and Neuroscience (IoPPN), King’s College London, London, UK; Ministry of Health Holdings Singapore, Singapore; Policy, Research and Evaluation Division, Ministry of Health, Singapore; Department of Neurology, National Neuroscience Institute (Singapore General Hospital Campus), Singapore

**Keywords:** Ethnicity, ischemic, stroke, young

## Abstract

**Objective:** There is a paucity of studies investigating the risk profile, and outcomes amongst young versus old stroke patients in Asian populations. Singapore is uniquely placed to contribute to the understanding of these risk factors, given our multi-ethnic population. Therefore, the aim of this study was to identify predictors of recurrence and outcome in stroke, comparing the young and the older stroke patients in Singapore.

**Methods:** This cohort study adhered to STROBE guidelines. Data were obtained from the Singapore Stroke Registry (SSR) from 2005 to 2016, to compare outcomes after first stroke among young (18-49 years) and among old stroke (≥50 years) patients. Outcome measures included recurrent stroke, acute myocardial infarction (AMI) and death from the above. Cox proportional hazards models were performed to determine risk factors for time to each outcome after first stroke, which included demographic and comorbidity variables.

**Results:** A total of 64,915 patients (6,705 young, and 58,210 old) were included in our analysis. After adjusting for gender, ethnicity, and comorbidities, old stroke patients have a greater hazard for recurrent stroke (HR = 1.20, 95%CI: 1.11-1.29), AMI (HR = 1.72, 95%CI: 1.52-1.94), and for recurrent stroke/AMI (HR = 1.36, 95%CI: 1.27-1.45). The old stroke patients were also more likely to die than younger stroke patients (HR = 2.62, 95%CI: 2.48-2.78). Amongst young stroke patients, males were at greater hazards for recurrent stroke (HR = 1.18, 95%CI: 1.00-1.38), AMI (HR = 1.40, 95%CI: 1.07-1.82), recurrent stroke/AMI (HR = 1.17, 95%CI: 1.02-1.36). However, males were at reduced hazards for death (HR = 0.87, 95%CI: 0.77-0.98). When analysing specifically young stroke patients, compared with the Chinese ethnicity, the Malay ethnicity was predisposed to recurrent stroke (HR = 1.36, 95%CI: 1.14-1.64). Compared with the Chinese ethnicity, both the Malay and Indian ethnicities were predisposed to AMI. The Malay ethnicity was at greater hazards for death (HR = 1.45, 95%CI: 1.27-1.66).

**Conclusions:** Non-Chinese ethnicities, especially Malay and Indian ethnicities experience poorer outcomes after first stroke. Further optimisation of risk factors targeting these high-priority populations are needed to achieve high quality care.

## Introduction

In contrast with the decreasing incidence of stroke in older adults, an increasing incidence and hospitalization rates for acute ischemic stroke in young adults have been identified.^1^ It has been estimated that the incidence of stroke in young adults increased up to 40% worldwide over the past decades, which has great impact because of high hospitalization costs an economic burden.^1^ Investigation of the prognosis of young patients who had stroke and the predictors of outcome and recurrence should be of paramount importance.

Few studies have evaluated inter-ethnic risk factor trends within a single co-localised Asian population.^2,3^ International research has provided evidence that ethnic minorities often have poorer access to best practice stroke care.^4^ Research on patient outcomes in different ethnic groups has produced conflicting results.^4-6^ Singapore is an economically-developed, small, tropical island city-state with a population of 5.5 million people. Of the 3.9 million residents (citizens and permanent residents), 74.3% are Chinese, 13.3% Malays, 9.1% Indians and 3.3% of other ethnicities (10).^7,8^ This multi-ethnic population facilitates assessment of ethnic differences and its associations with outcomes. Therefore, the aim of this study to identify predictors of recurrence and outcome in stroke within the Asian population in Singapore. We also aimed to determine if there were any specific populations at high risk of recurrent events within young stroke patients.

## Methods

### Data collection

Data were obtained from the Singapore Stroke Registry (SSR) from 2005 to 2016, to compare outcomes after first stroke among young (18-49 years) and old stroke (≥50 years) patients. This cohort study adhered to the Strengthening the Reporting of Observational studies in Epidemiology (STROBE) guidelines.^9^ Ethics approval was not required as this study was registered as a surveillance study.

The SSR is an active nationwide prospective registry collecting data of stroke cases from all public hospitals in Singapore.^8^ It measures the incidences of stroke, separated by first from recurrent stroke, determines risk factors and subtypes, assessed prognosis including early and late case-fatality, and uncovers inter-ethnic variations of stroke patterns. The aforementioned data were then matched to two databases using the patients’ unique National Registration Identity Card number for outcomes and pre-existing events: (i) the National Death Registry and (ii) the Singapore Myocardial Infarct Registry.

The collected patient variables included: age, gender, smoking history, and comorbidities (hypertension, hyperlipidaemia, diabetes mellitus, and ischaemic heart disease), and outcomes such as stroke recurrence, acute myocardial infarction (AMI) and death.

### Exposures and outcomes measures

Patients were separated into two age groups: <50 and ≥50 years. The primary outcome measure was recurrent stroke. The secondary outcomes were AMI, a combined endpoint of stroke and AMI, and death. Patients are followed up from date of first stroke until 31 December 2016 or date of death, whichever is earlier.

### Statistical analysis

Comparisons of categorical variables were performed using the Pearson chi-squared test. Cox proportional hazard regressions were used to identify independent risk factors for time to each outcome after first stroke, which included demographic and comorbidity variables. Analyses were also performed separately in young stroke patients. Tests for proportional hazard assumption were done using log-log plots and comparison of Kaplan-Meier observed survival curves with the Cox predicted curves.

Data management and statistical analyses were performed using STATA/SE 16.0 (StataCorp LLC, USA). P-values less than 0.05 were considered statistically significant.

## Results

### Baseline characteristics

A total of 64,915 patients were included in our analysis. There were 6,705 patients aged 18-49 years and 58,210 aged ≥50 years at first stroke. Table 1 compares baseline patient characteristics between the two age groups. The most common comorbidities were hypertension, hyperlipidaemia, diabetes mellitus and ischemic heart disease (IHD).

**Table 1.** Baseline characteristics of the study population.

|  | **18-49 years**<br>**N=6,705** | **≥50 years**<br>**N=58,210** | ***p* value** |
| --- | --- | --- | --- |
| Gender |  |  |  |
| Female | 2,344 (35%) | 26,069 (45%) | <0.001 |
| Male | 4,361 (65%) | 32,141 (55%) |  |
| Ethnicity |  |  |  |
| Chinese | 4,508 (67%) | 45,420 (78%) | <0.001 |
| Malay | 1,044 (16%) | 6,905 (12%) |  |
| Indian | 631 (9%) | 3,507 (6%) |  |
| Others | 522 (8%) | 2,378 (4%) |  |
| Comorbidities |  |  |  |
| TIA | 135 (2%) | 2,063 (4%) | <0.001 |
| Hypertension | 3,532 (53%) | 44,200 (76%) | <0.001 |
| DM | 1,446 (22%) | 21,408 (37%) | <0.001 |
| Ischemic heart disease | 462 (7%) | 13,131 (23%) | <0.001 |
| Atrial fibrillation | 124 (2%) | 6,693 (12%) | <0.001 |
| Valve heart disease | 136 (2%) | 1,237 (2%) | 0.602 |
| Peripheral vascular disease | 97 (1%) | 1,772 (3%) | <0.001 |
| Hyperlipidaemia | 1,927 (29%) | 31,059 (53%) | <0.001 |
| Smoking | 634 (9%) | 4,775 (8%) | <0.001 |
| Outcomes |  |  |  |
| AMI after stroke | 341 (5%) | 5,283 (9%) | <0.001 |
| Recurrent stroke | 751 (11%) | 7,255 (12%) | 0.003 |
| AMI/Recurrent stroke | 1,015 (15%) | 11,509 (20%) | <0.001 |
| Death | 1,153 (17%) | 24,698 (42%) | <0.001 |

All categorical data presented as n (%). *p* values are from $\chi^2$ tests.
AMI = Acute Myocardial Infarction; DM = Diabetes Mellitus; TIA = Transient Ischemic Attack

### Overall risk of outcomes after first stroke amongst old and young patients

After adjusting for gender, ethnicity, and comorbidities, older stroke patients have a greater risk for recurrent stroke (HR = 1.21, 95%CI: 1.12-1.30, p < 0.001), AMI (HR = 1.73, 95%CI: 1.54-1.95, p < 0.001), and for recurrent stroke/AMI (HR = 1.37, 95%CI: 1.28-1.46, p < 0.001). The old stroke patients were also more likely to die than younger stroke patients (HR = 2.49, 95%CI: 2.34-2.64, p < 0.001) (Table 2).

**Table 2.**
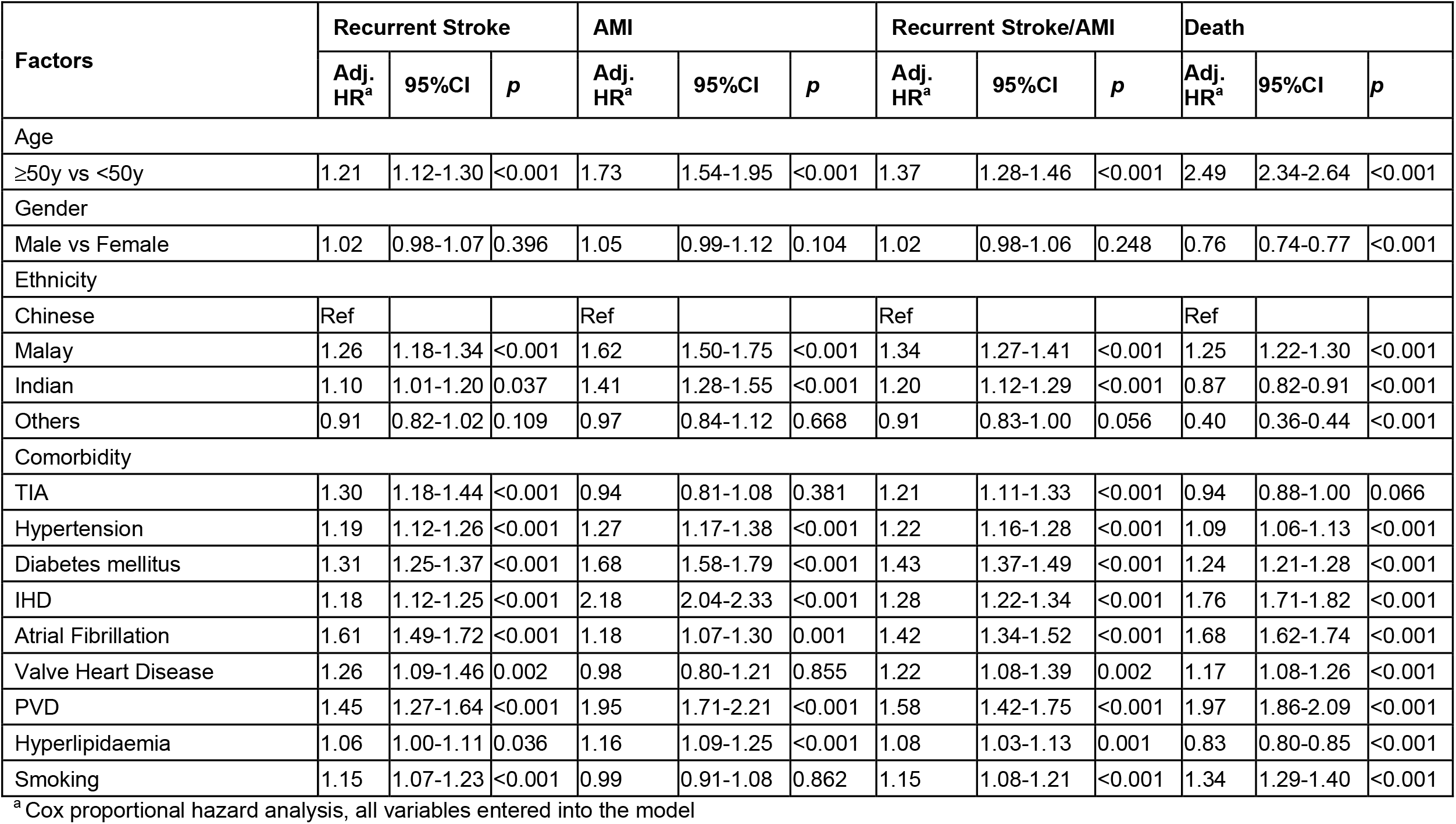
Risk of outcomes after first stroke.

Compared with the Chinese ethnicity, the Malay ethnicity was predisposed to recurrent stroke (HR = 1.26, 95%CI: 1.18-1.34, p < 0.001). Compared with the Chinese ethnicity, both the Malay and Indian ethnicities were predisposed to AMI (HR = 1.62, 95%CI: 1.50-1.75, p < 0.001, and HR = 1.41, 95%CI: 1.28-1.55, p < 0.001 respectively). Compared with the Chinese ethnicity, the Malay ethnicity was at greater hazards for death (HR = 1.25, 95%CI: 1.22-1.30, p < 0.001), whilst the Indian ethnicity was a lower hazards for death (HR = 0.87, 95%CI: 0.82-0.91, p < 0.001).

### Risk of outcomes after first stroke amongst young patients

Amongst young stroke patients, males were at greater hazards for recurrent stroke (HR = 1.18, 95%CI: 1.01-1.39, p = 0.043), AMI (HR = 1.41, 95%CI: 1.08-1.83, p = 0.012), recurrent stroke/AMI (HR = 1.18, 95%CI: 1.02-1.36, p = 0.026). However, males were at reduced hazards for death (HR = 0.78, 95%CI: 0.69-0.89, p < 0.001) (Table 3).

**Table 3.** Risk of outcomes after first stroke amongst young patients.

| Factors | Recurrent Stroke |  |  | AMI |  |  | Recurrent Stroke /AMI |  |  | Death |  |  |
| --- | --- | --- | --- | --- | --- | --- | --- | --- | --- | --- | --- | --- |
|  | Adj. HR<sup>a</sup> | 95%CI | *p* | Adj. HR<sup>a</sup> | 95%CI | *p* | Adj. HR<sup>a</sup> | 95%CI | *p* | Adj. HR<sup>a</sup> | 95%CI | *p* |
| Gender |  |  |  |  |  |  |  |  |  |  |  |  |
| Male vs Female | 1.18 | 1.01-1.39 | 0.043 | 1.41 | 1.08-1.83 | 0.012 | 1.18 | 1.02-1.36 | 0.026 | 0.78 | 0.69-0.89 | <0.001 |
| Ethnicity |  |  |  |  |  |  |  |  |  |  |  |  |
| Chinese | Ref |  |  | Ref |  |  | Ref |  |  | Ref |  |  |
| Malay | 1.37 | 1.14-1.65 | 0.001 | 2.45 | 1.87-3.22 | <0.001 | 1.59 | 1.35-1.87 | <0.001 | 1.43 | 1.24-1.66 | <0.001 |
| Indian | 0.97 | 0.76-1.24 | 0.783 | 1.96 | 1.41-2.72 | <0.001 | 1.10 | 0.89-1.36 | 0.382 | 0.99 | 0.81-1.22 | 0.955 |
| Others | 0.75 | 0.55-1.02 | 0.063 | 0.98 | 0.60-1.60 | 0.935 | 0.75 | 0.57-0.99 | 0.045 | 0.32 | 0.22-0.47 | <0.001 |
| Comorbidity |  |  |  |  |  |  |  |  |  |  |  |  |
| TIA | 1.65 | 1.15-2.36 | 0.007 | 1.67 | 0.95-2.94 | 0.075 | 1.56 | 1.12-2.18 | 0.009 | 0.62 | 0.39-0.99 | 0.047 |
| Hypertension | 1.52 | 1.28-1.80 | <0.001 | 1.52 | 1.15-2.02 | 0.004 | 1.55 | 1.33-1.80 | <0.001 | 1.09 | 0.95-1.25 | 0.205 |
| DM | 1.86 | 1.56-2.21 | <0.001 | 2.95 | 2.27-3.84 | <0.001 | 2.11 | 1.81-2.46 | <0.001 | 1.57 | 1.35-1.82 | <0.001 |
| IHD | 1.27 | 1.00-1.62 | 0.049 | 2.43 | 1.78-3.31 | <0.001 | 1.26 | 1.02-1.57 | 0.035 | 1.66 | 1.37-2.03 | <0.001 |
| Atrial Fibrillation | 0.88 | 0.50-1.52 | 0.641 | 1.11 | 0.48-2.59 | 0.805 | 0.80 | 0.47-1.36 | 0.405 | 1.40 | 0.98-1.98 | 0.063 |
| Valve Heart Disease | 1.53 | 0.92-2.52 | 0.098 | 0.36 | 0.08-1.35 | 0.123 | 1.35 | 0.84-2.16 | 0.211 | 1.97 | 1.41-2.76 | <0.001 |
| PVD | 1.78 | 1.15-2.76 | 0.010 | 1.36 | 0.71-2.61 | 0.354 | 1.58 | 1.05-2.38 | 0.030 | 2.10 | 1.53-2.89 | <0.001 |
| Hyperlipidaemia | 1.14 | 0.95-1.35 | 0.158 | 1.04 | 0.79-1.37 | 0.775 | 1.10 | 0.94-1.28 | 0.247 | 0.91 | 0.78-1.06 | 0.222 |
| Smoking | 1.20 | 0.98-1.46 | 0.079 | 0.96 | 0.71-1.30 | 0.768 | 1.23 | 1.03-1.46 | 0.022 | 1.26 | 1.05-1.51 | 0.012 |

<sup>a</sup> Cox proportional hazard analysis, all variables entered into the model

In younger patients, compared with the Chinese ethnicity, the Malay ethnicity was predisposed to recurrent stroke (HR = 1.37, 95%CI: 1.14-1.65, p < 0.001). Compared with the Chinese ethnicity, both the Malay and Indian ethnicities were predisposed to AMI (HR = 2.45, 95%CI: 1.87-3.22, p < 0.001, and HR = 1.96, 95%CI: 1.41-2.72, p < 0.001 respectively). The Malay ethnicity was at greater hazards for death (HR = 1.43, 95%CI: 1.24-1.66, p < 0.001).

## Discussion

In order to improve stroke outcomes and to achieve the impact goals set by the American Heart Association to reduce stroke death, future epidemiologic studies that investigate and identify the most efficient and effective clinical approaches to both prevention and treatment are important. This study presents new evidence on the association between ethnicity and post-stroke outcomes from a prospective nationwide study of all admitted patients with stroke. This study demonstrates that older stroke patients were more likely to suffer from recurrent stroke, AMI and death. Within the young stroke population specifically, males were predisposed to recurrent stroke and AMI but were protected against death. Within our multi-ethnic population, both the Malay and Indian ethnicities were predisposed to AMI, whilst the Malay ethnicity was particularly vulnerable to recurrent stroke and death.

We postulate that the findings of increased stroke recurrence in the elderly group to the increased incidence of traditional risk factors, such as hypertension, in this age group.^10^ In addition, previous studies have shown that, compared with the elderly, young patients who had ischaemic stroke have lower National Institutes of Health Stroke Scale (NIHSS) scores and hence milder stroke severity at the time of their hospital admission.^3,11^ This is suggested that young patients were more active in seeking medical attention; thus, they had low NIHSS scores at the time of their hospital admission and subsequently more favorable post-stroke outcomes.

Despite adjusting the analysis for stroke risk factors and baseline disability, it is possible that a degree of residual confounding could explain some of the disparities between the ethnic groups. Some of the outcome differences are probably still influenced by poorer underlying health status and we should not dismiss the importance of risk factor management. The higher prevalence of known stroke risk factors among the Malay and Indian ethnic patients in this cohort may have been attributable for the larger proportion of stroke recurrence and poor outcomes. Studies have shown that amongst the South Asian community, hypertension is more prevalent and is a potent risk factor for stroke recurrence, distantly trailed by diabetes mellitus and cardiac diseases.^12,13^ Stroke remains a major health-care problem in Singapore and is the fourth leading cause of death, highest among Malay women.^14^ Our findings support this notion as we found that female gender and the Malay ethnicity was particularly vulnerable to death.

These findings support the need for genetic and metabolomics studies that may lead to an understanding of the mechanisms underlying the ethnic variations. The further examination of the interaction between ethnicity and stroke risk factors can provide data that will enable clinicians to predict stroke outcomes after an acute stroke admission and to act on that knowledge accordingly.

### Limitations

Despite adjusting results for established strong predictors of stroke outcomes and differences in baseline characteristics, residual confounding cannot be excluded. Importantly, the SSR does not classify AIS cases into stroke aetiology subtypes, limiting the analysis of specific reasons behind our analyses. However, as longitudinal data on post-stroke outcomes is scarce, this study still provides valuable insight and encourages further research efforts in this domain. Additionally, the merits of this study include the large database of patients giving excellent representation and minimal risk of selection bias whilst conferring statistical power to detect disparities in outcomes. Furthermore, the study of inter-ethnic risk factor trends within a single multi-ethnic composition, uniquely to Singapore, is another strength of this study.

## Conclusion

Compared with the Chinese ethnicity, young stroke especially in Malay and Indian ethnicities experience poorer outcomes after first stroke. Further optimisation of risk factors targeting these high-priority populations are needed to achieve high quality care.

## Data Availability

Data are available upon request from the corresponding author.

